# Association between nursing home crowding and outbreak-associated respiratory infection and death prior to the COVID-19 pandemic between 2014 and 2019 in Ontario, Canada

**DOI:** 10.1101/2022.07.06.22277066

**Authors:** Pamela Leece, Michael Whelan, Andrew P. Costa, Nick Daneman, Jennie Johnstone, Allison McGeer, Paula Rochon, Kevin L. Schwartz, Kevin A. Brown

## Abstract

**Importance:** Resident crowding in nursing homes is associated with larger SARS-CoV-2 outbreaks. However, this association has not been previously documented for non-SARS-CoV-2 respiratory infections.

**Objective:** We sought to measure the association between nursing home crowding and respiratory infections in Ontario nursing homes prior to the COVID-19 pandemic.

**Design, Setting, and Participants:** We conducted a retrospective cohort study of nursing home residents in Ontario, Canada over a five-year period prior to the COVID-19 pandemic, between September 2014 and August 2019.

**Exposure:** Using administrative data, we estimated the crowding index equal to the mean number of residents per bedroom and bathroom (residents / [0.5*bedrooms+0.5*bathrooms]).

**Outcomes:** The incidence of outbreak-associated infections and mortality per 100 nursing home residents per year. We also examined infection and mortality outcomes for outbreaks due to 7 specific pathogens: coronaviruses (OC43, 229E, NL63, HKU1), influenza A, influenza B, human metapneumovirus, parainfluenza virus, respiratory syncytial virus, rhinovirus/enterovirus.

**Results:** There was one or more respiratory outbreak in 93.9% (588/626) nursing homes in Ontario. There were 4,921 outbreaks involving 64,829 cases of respiratory infection, and 1,969 deaths. Outbreaks attributable to a single identified pathogen were principally caused by influenza A (29%), rhinovirus (11.7%), influenza B (8.1%), and respiratory syncytial virus (6.1%). Among homes, 42.7% (251/588) homes had a high crowding index (≥ 2.0). After adjustment, more crowded homes had higher outbreak-associated respiratory infection incidence (aRR 1.89; 95% 1.64-2.18) and mortality incidence (aRR 2.28; 95% 1.84-2.84). More crowded homes had higher adjusted estimates of the incidence of infection and mortality for each of the 7 respiratory pathogens examined.

**Conclusions and Relevance:** Residents of crowded nursing homes experienced more respiratory-outbreak infections and mortality due to influenza and other non-SARS-CoV-2 respiratory pathogens. Decreasing crowding in nursing homes is an important patient safety target beyond the COVID-19 pandemic.

## Introduction

Even prior to the COVID-19 pandemic, nursing homes were severely impacted by outbreaks of respiratory infections, particularly influenza.^1,2^ This is because residents of nursing homes are more exposed to infection due to the congregate nature of nursing homes, and because residents are more likely to develop severe disease due to their advanced age, comorbidities and a high degree of frailty.^3^ Several design features of nursing homes could aggravate or mitigate the potential for transmission in a home; one important feature is the degree of crowding, which can be defined in terms of occupants per square foot, per room, or per bedroom. For nursing homes, one simple approach for measuring the degree of crowding is the average number of residents per occupied bedroom and bathroom across a home.^4^ Crowding defined in this way has been associated with nursing home SARS-CoV-2 infection rates, and mortality during the COVID-19 pandemic.^4^ There is substantial evidence that crowding is associated with transmission of a number of infections, particularly tuberculosis, in private dwellings.^5^ Evidence on whether crowding is also a risk factor for non-SARS-CoV-2 respiratory infections could influence the extension of crowding restrictions beyond the pandemic period, as well as the design standards for the construction of new nursing homes.^6^ We undertook this study to examine the association between crowding and nursing home outbreak-associated respiratory infection incidence and related mortality in Ontario nursing homes, prior to the COVID-19 pandemic.

## Methods

### Population, Study Design, and Data Sources

We conducted a retrospective cohort study of nursing homes in Ontario, between September 2014 and August 2019. The research ethics board of Public Health Ontario reviewed and approved this study. Data on respiratory infection outbreaks were retrieved from the integrated Public Health Information System of Ontario. Under the Ontario Public Health Standards, respiratory infection outbreaks in institutions and public hospitals require provincial reporting. Respiratory infection outbreaks in Ontario are defined as either two cases of acute respiratory infection (new or worsening cough or shortness of breath) within 48 hours in a nursing home with an epidemiologic link (e.g., unit or floor), including one laboratory-confirmed case, or three cases regardless of laboratory confirmation.^7^ Nursing home characteristics were obtained from the Ministry of Long-Term Care, inspections branch, extracted on January 2020; some nursing homes in Ontario are not administered by the Ministry of Long-Term Care and could not be included, as well as homes that closed before January 2020.

### Crowding index

The exposure of interest was the nursing home crowding index, which we defined as the average number of residents per bedroom and bathroom across the home (i.e. residents / (0.5*bedrooms + 0.5*bathrooms).^4^ Only bathrooms intended exclusively for resident use, and located in sleeping quarters, are included in the calculation. A home composed exclusively of single-bed rooms with private bathrooms would have a crowding index of 1, while a home composed exclusively of 4-bed rooms would have a crowding index of 4. If half of residents resided in single-bed rooms with private bathrooms, and the other half in 4-bed rooms each with its own shared bathroom, the crowding index would be 2.5. Because we lacked direct measurements of nursing home crowding, we used a validated approach to estimate the crowding index of Ontario nursing homes.

Specifically, the Ontario Ministry of Long Term Care provided information on the distribution of new beds and type A beds (meeting the 1999 design standard) versus type B, C, or D beds (not meeting the 1999 design standard), and the class of the bed (private, semi-private, or basic). New design standards stipulate that “private” beds are located in 1-bed rooms with private bathroom (crowding weight=1), “semi-private” beds are located in 1-bed rooms with a shared bathroom (crowding weight=1.5) and that “basic” rooms are located in 2-bed rooms (crowding weight=2). In older homes, standards indicate that “private” rooms are 1-bed rooms that can have shared bathrooms (crowding weight=1.5), and that “semi-private” rooms are 2-bed rooms (crowding weight=2). There was substantial variability in the number of residents per room among “basic” beds in older homes, and which could have up to 5 beds per room. We asked the Ministry of Long Term Care to use survey data from 2022 to determine the average number of persons per room among “basic” beds in older homes; this was determined to be 2.04 for municipal homes, 2.50 for non-profit homes, and 3.20 among for profit homes; these figures were used as weights applied to the proportion of basic beds in old homes. In prior work,^4^ we originally used weights of 4 for all basic beds in old homes; our new validated approach was strongly correlated with the original approach (Pearson correlation = 0.96) but yielded lower overall estimates of crowding (median validated crowding index = 1.7, IQR: 1.4, 2.5; median original crowding index = 1.9, IQR: 1.4, 2.9) and lower estimates of the overall number of beds in shared rooms with 3 or more beds (8,402 versus 15,812).

### Outcomes

Our primary outcome was the incidence of outbreak-associated respiratory infections per 100 nursing home residents per year recorded as part of a nursing home respiratory outbreak. Secondary outcomes included: (1) incidence of outbreak-associated deaths per 100 nursing home residents per year, where only cases that died as a result of the infection are included (as per the outbreak investigator and/or the most responsible physician),^8^ (2) outbreak frequency per year, and (3) outbreak size as a proportion of the number of residents in the home. We also examined incidence of infections and deaths for specific pathogens, classified into 10 categories as follows: coronaviruses (OC43, 229E, NL63, HKU1), influenza A, influenza B, human metapneumovirus, parainfluenza virus, respiratory syncytial virus, rhinovirus/enterovirus, other (adenovirus), multiple agents, and unidentified. For descriptive purposes, we also examined the case fatality rate (deaths divided by infections).

### Nursing home characteristics

We included data on nursing home ownership (private for-profit entity, private non-profit, or owned by a municipality), nursing home size (<100 beds, ≥100 beds), nursing home health region (N=6: East, Central-East, Toronto, Central-West, South-West, North).

### Resident characteristics

Aggregate characteristics of nursing home residents were obtained from the Resident Assessment Instrument Minimum Data, extracted for January 2020.^9^ Variables included were mean resident age, female sex (%), diabetes (%), and the mean of the activities of daily living (ADL) 6-point scale (0, independent to 6, total dependence).

### Statistical analyses

To derive p-values for comparing characteristics of high crowding index homes (>=2) versus low crowding index homes (<2), we used logistic regression. We used negative binomial regression to model the incidence of outbreak-associated respiratory infections and deaths occurring within a home with an offset of the number of beds in the home. To model the outbreak frequency, we used negative binomial regression without an offset. To model outbreak size, we used logistic regression, with a random effect for home, since there could be multiple outbreaks per home. All unadjusted models included just the continuous crowding index as the only covariate. All adjusted models included the crowding index, in addition to home characteristics (size, ownership, region) and aggregate resident characteristics (age, female sex, diabetes, and ADL hierarchy). Crowding index effects were scaled to accurately reflect the comparison between the average home with crowding index ≥ 2 versus the average home with crowding index <2 by scaling effect estimates to the difference in mean crowding index between these two groups.

Further, we ran specific models for incidence of infections and deaths for each of 10 pathogen groups described above. We recombined pathogen specific models using random effects meta-analysis, and reported the combined meta-analytic estimate, as well as the degree of heterogeneity between the pathogen specific estimates, using the Higgins (I^2^) statistic.^10^ We used simulations to estimate the annual incidence of respiratory infections and deaths for less crowded conditions. We produced estimates for scenarios where all 4-bed rooms were replaced with 2-bed rooms, and 2- and 4-bed rooms were replaced with 1-bed rooms (over 95% of residents are housed in 1, 2, or 4-bed rooms in Ontario).

Statistical analysis was conducted using R version 4.1.0; generalized additive models were fit with the mgcv package, while random-effects meta-analysis used the metafor package. P-values were based on 2-sided testing.

## Results

During the five-year study period, there were 5,107 nursing home respiratory outbreaks recorded in Ontario, of which 4,921 (96.4%) were included in the study, from across 588 of 626 (93.9%) of nursing homes in the province. Among these outbreaks, there was a total of 64,829 cases of acute respiratory infection (17.2% of residents per year), and 1,969 deaths (0.52% of residents per year; 3.04% case fatality rate; Table 1).

**Table 1.** Characteristics and outcomes associated with low and high crowding index homes.

| Characteristic | Home crowding index |  |  | P-value |
| --- | --- | --- | --- | --- |
|  | All homes | Low (< 2.0) | High (>= 2.0) |  |
| Homes (N, %) | 588 (100) | 337 (100) | 251 (100) |  |
| **Facility characteristics** |  |  |  |  |
| Crowding index (median, IQR) | 1.9 (1.4, 2.5) | 1.5 (1.4, 1.6) | 2.5 (2.4, 2.6) | < 0.001 |
| Ownership (N, %) |  |  |  |  |
| Municipal | 96 (16.3) | 95 (28.2) | 1 (0.4) | < 0.001 |
| Private, for-profit | 343 (58.3) | 123 (36.5) | 220 (87.6) | < 0.001 |
| Private, non-profit | 149 (25.3) | 119 (35.3) | 30 (12.0) | < 0.001 |
| Size of home |  |  |  |  |
| Number of beds (median, IQR) | 130 (75, 160) | 149 (101, 175) | 105 (60, 126) | < 0.001 |
| >= 100 beds (N, %) | 365 (62.1) | 262 (77.7) | 103 (41.0) | < 0.001 |
| Occupancy rate (median, IQR) | 98.2 (97.5, 100) | 98.5 (97.9, 99.6) | 97.8 (96.7, 100.0) | < 0.001 |
| **Resident characteristics (median, IQR)** |  |  |  |  |
| Mean age | 83.5 (82.2, 85.1) | 84.5 (83.4, 85.7) | 82.1 (80.6, 84.0) | < 0.001 |
| Percent Female | 68.6 (64.2, 73.3) | 70.2 (66.7, 74.0) | 66.4 (60.7, 72.6) | < 0.001 |
| Diabetes | 28.0 (23.5, 31.9) | 26.3 (22.8, 29.8) | 30.2 (25.6, 34.8) | < 0.001 |
| ADL 6-point scale* | 3.9 (3.6, 4.1) | 3.9 (3.6, 4.2) | 3.8 (3.6, 4.1) | 0.13 |
| **Outcomes** |  |  |  |  |
| Resident Infections<br>*N (per 100 resident-years)* | 64,829 (17.2) | 33,485 (13.5) | 31,344 (24.2) | < 0.001 |
| Resident Deaths<br>*N (N per 100 resident-years)* | 1,969 (0.5) | 1,007 (0.4) | 962 (0.7) | < 0.001 |
| Case Fatality Ratio<br>*Deaths per 100 infections* | 3.04 | 3.01 | 3.07 | 0.15 |
| Outbreak Frequency<br>*N (N per home-year)* | 4,921 (1.7) | 2,844 (1.7) | 2,077 (1.7) | 0.72 |
| Mean Outbreak Size<br>*N (% of home residents)* | 13.2 (11.5) | 11.8 (8.2) | 15.1 (16.0) | < 0.001 |

\* Activities of Daily Living (0=independent, 6=total dependence)

### Virologic findings

Influenza A was the most common pathogen identified among outbreak associated cases, and had the highest incidence rate (Table 2, 22,159; 5.9% of residents per year) followed by rhinovirus (7,316, 1.9% of residents per year) and influenza B (5,289, 1.4% of residents per year). Influenza A, with the highest case fatality (4.2%) caused the most outbreak-associated deaths (937, 0.25% of residents per year), followed by influenza B (233, 0.06% of residents per year), and respiratory syncytial virus (116, 0.03% of residents per year).

**Table 2.** Viral agents identified for outbreak-associated viral respiratory infections in Ontario, 2014-2019.

|  | Resident<br>Infections | Resident<br>Deaths | Case Fatality<br>Ratio | Outbreak<br>Frequency | Mean Outbreak<br>Size |
| --- | --- | --- | --- | --- | --- |
|  | *N (N per 100*<br>*resident-years)* | *N (N per 100*<br>*resident-years)* | *Deaths per*<br>*100 infections* | *N (N per home*<br>*per year)* | *N (% of home*<br>*residents)* |
| Single Agent |  |  |  |  |  |
| Coronaviruses* | 2,385 (0.6) | 25 (0.01) | 1.05 | 197 (0.1) | 12.1 (10.8) |
| Human metapneumovirus | 1,969 (0.5) | 59 (0.02) | 3.00 | 162 (0.1) | 12.2 (12.1) |
| Human parainfluenza<br>viruses** | 2,829 (0.8) | 80 (0.02) | 2.83 | 232 (0.1) | 12.2 (10.8) |
| Influenza A | 22,159 (5.9) | 937 (0.25) | 4.23 | 1,414 (0.5) | 15.7 (12.8) |
| Influenza B | 5,289 (1.4) | 233 (0.06) | 4.41 | 391 (0.1) | 13.5 (11.8) |
| Respiratory syncytial virus | 3,954 (1.0) | 116 (0.03) | 2.93 | 309 (0.1) | 12.8 (11.7) |
| Rhinovirus/enterovirus | 7,316 (1.9) | 108 (0.03) | 1.48 | 591 (0.2) | 12.4 (10.9) |
| Other | 772 (0.2) | 36 (0.01) | 4.66 | 57 (0.0) | 13.5 (13.6) |
| Multiple Agents | 4,501 (1.2) | 162 (0.04) | 3.60 | 280 (0.1) | 16.1 (13.6) |
| Unidentified | 13,655 (3.6) | 213 (0.06) | 1.56 | 1,288 (0.4) | 10.6 (9.8) |

\* Coronavirus strains OC43, 229E, NL63, HKU1 were tested; \*\* Human parainfluenza strains 1-4 were tested.

### Crowding index

Crowded homes (crowding index ≥2) represented 42.6% (251) of homes included in this study. High crowding index homes were more likely to be private for-profit homes (87.6%) compared to low crowding homes (36.5%, p < 0.001). High crowding index homes also tended to be smaller (median 105 beds), compared to low crowding homes (median = 149 beds, p < 0.001)

### Association between crowding and respiratory infections

Compared to low crowding index homes, high crowding index nursing homes had a higher incidence rate of outbreak-associated infections per 100 residents per year (24.2% vs. 13.5%, p < 0.001) and deaths (0.74% vs. 0.41%, p < 0.001), and outbreak size (16.0% vs. 8.2% of residents infected per outbreak, p < 0.01); frequency of outbreaks was similar between the 2 groups.

Unadjusted and adjusted models (Table 3) corroborated that nursing homes with high crowding index had higher incidence of outbreak-associated infections (aRR 1.89; 95% 1.64, 2.18) and deaths (aRR 2.28; 95% 1.84, 2.84), outbreak frequency (aRR 1.32; 95% 1.15, 1.51) and outbreak size (aRR 1.40; 95% 1.28, 1.54) when compared to nursing homes with low crowding index. Further, there was a positive dose-response association between crowding and incidence of infections and deaths (Figure 1).

**Table 3.** Unadjusted and adjusted association of crowding index with outbreak-associated respiratory

| Characteristic | Infection<br>Incidence | Death<br>Incidence | Outbreak<br>Frequency | Outbreak<br>Size |
| --- | --- | --- | --- | --- |
|  | RR (95% CI) | RR (95% CI) | RR (95% CI) | RR (95% CI) |
| **Unadjusted Analysis** |  |  |  |  |
| Crowding index $\geq 2.0$ | 1.94 (1.75, 2.14) | 1.98 (1.70, 2.31) | 1.02 (0.92, 1.13) | 1.81 (1.67, 1.96) |
| **Adjusted Analysis** |  |  |  |  |
| Crowding index $\geq 2.0$ | 1.89 (1.64, 2.18) | 2.28 (1.84, 2.84) | 1.32 (1.15, 1.51) | 1.40 (1.28, 1.54) |

\* Adjusted for ownership type (municipal, private for-profit, private non-profit), size of home, mean age of residents, proportion female, proportion with diabetes, and the mean activities of daily living score, and health region of the nursing home (N=6 regions); Note: RR, risk ratio.

**Figure 1.**
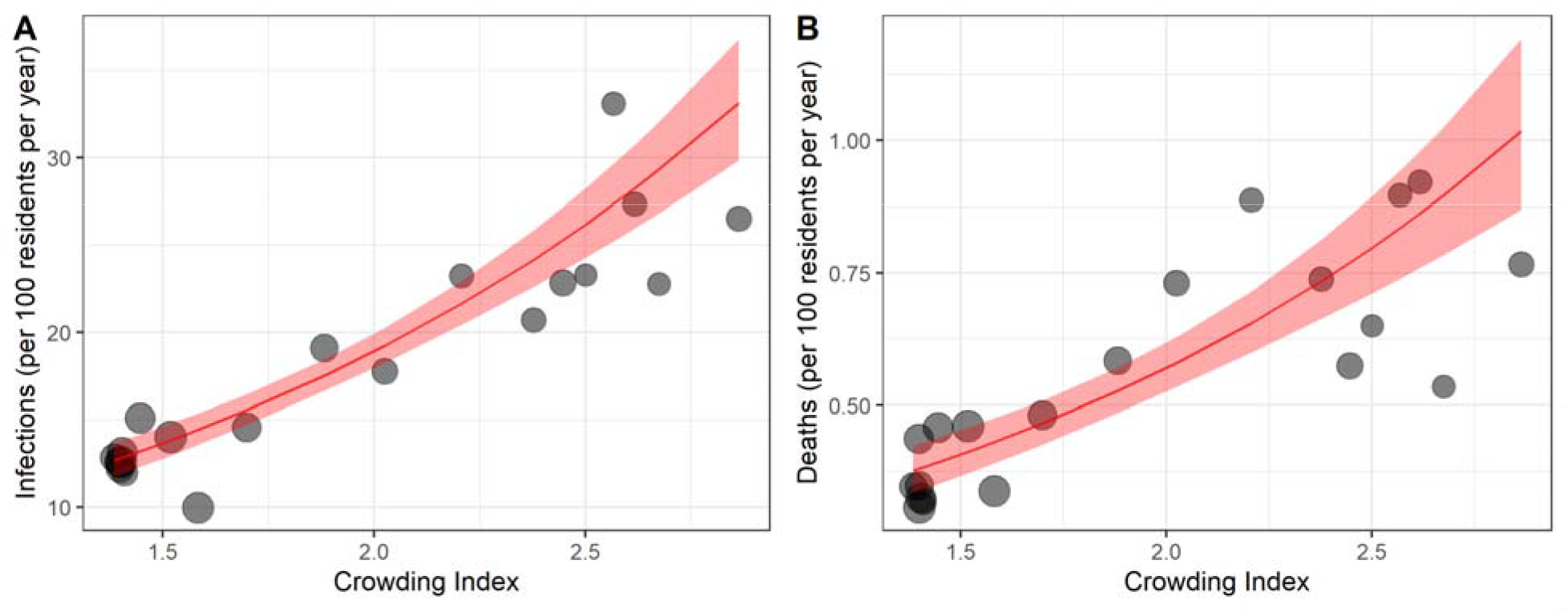
Association between nursing home crowding index and incidence of outbreak-associated respiratory infections (panel A) and deaths (panel B). For visualizing trends in the nursing home data, homes were combined into 20 equal-sized groups of 29-30 homes according to crowding index (black points). The red line reflects the negative binomial model-based association with the crowding index, along with the 95% confidence band. Each 1-point increase in the crowding index was associated with a 1.92-fold increase in incidence of infection (95%CI: 1.74, 2.11, Figure 1), and a 1.96-fold increase in respiratory infection associated mortality (95%CI: 1.68, 2.28).

We also fitted the incidence of infections and deaths outcomes separately for each respiratory pathogen, adjusting for nursing home size, ownership type, and mean age (adjustment for all caused convergence issues for some models). For incidence of infections, the associations between crowding and specific infectious agents (Figure 2) ranged from 1.40 (95%CI: 0.68, 2.87) for coronaviruses, to 2.92 (95%CI: 1.88, 4.54) for rhinovirus; the association for influenza A virus was 1.52 (95%CI: 1.27, 1.82). Random-effects meta-analysis across pathogens for incidence of infection was similar to the primary analysis (RR=1.84, 95%CI: 1.54, 2.19), and revealed some heterogeneity (I^2^ = 30%). For incidence of deaths, patterns were similar, (RR=2.13, 95%CI: 1.70, 2.65, I^2^ = 19%).

**Figure 2.**
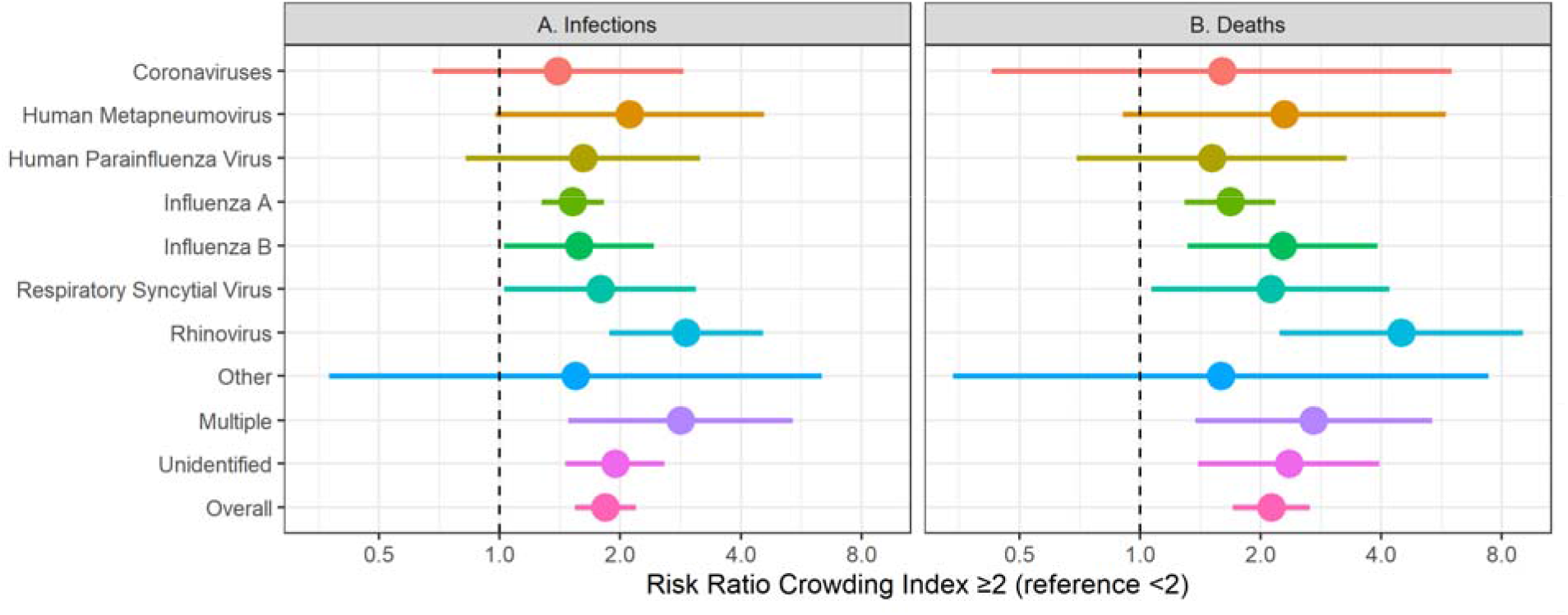
Adjusted association between nursing home crowding index and respiratory pathogen-specific outbreak associated (1) infections (left panel) and (2) deaths (right panel). Points represent the risk ratios (comparing homes with crowding index <2.0 versus homes with crowding index ≥2.0) and line ranges represent the corresponding 95% confidence intervals. For each pathogen, the adjustment models included nursing home size, ownership type, and mean resident age. The overall estimate is based on random effects meta-analysis. * Coronavirus strains OC43, 229E, NL63, HKU1 tested; ** Parainfluenza strains 1-4 tested.

### Simulations

We used simulations to examine the potential impacts of an intervention to change the number of persons residing in shared rooms. We found that a cap on the capacity of rooms at 2-beds would have reduced infections over the 5-year period from 64,829 to 53,650 (95% CI: 50,571 to 57,425, reduction of 11,179, 95%CI: 7,404, 14,258), and a nursing home system with exclusively single-bedded rooms (with private bathrooms) would have experienced 36,222 (95%CI: 32,027 to 41,432, reduction of 28,607, 95%CI: 23,397, 28,607). A cap on the capacity of rooms at 2-beds would have reduced outbreak-associated deaths from 1,969 to 1,553 (95% CI: 1,398 to 1,698, reduction of 416, 95%CI: 271, 571), and a nursing home system with exclusively single-bedded rooms (with private bathrooms) would have experienced 949 outbreak-associated deaths (95%CI: 776 to 1,149), a reduction of 1020 deaths (95%CI: 820, 1193).

## Discussion

Over a five-year period in Ontario, there were 64,829 cases and 1,969 deaths related to nursing home respiratory outbreaks involving 588 nursing homes. Crowded nursing homes had greater incidence of outbreak-related acute respiratory infections and associated mortality, as well as respiratory infection outbreak frequency and size. Crowding was positively associated with increased incidence for all 7 of the respiratory viruses examined. Simulations estimated that deaths associated with respiratory outbreaks would be reduced by over 50% if nursing home occupancy was reduced to 1 person per room.

There is relatively little systematic data on the presence of shared rooms, so-called ‘ward rooms’, in nursing homes. While national statistics agencies in several countries collect measures of crowding for residents of private dwellings (e.g., the Canadian National Occupancy Standard), the same measures are usually not collected or reported for residents of nursing homes. In the absence of systematic data from the United States, a survey from 40 randomly selected nursing homes across 5 US states (California, Florida, Minnesota, New Jersey, and New York) indicated that 71% of nursing home residents lived in shared rooms, and 28% of nursing home residents shared a bathroom between 4 or more residents; of note, all homes with private rooms were non-profit facilities.^11^ Similarly, there is no systematic, country wide data on crowding in nursing homes in Canada. In this cohort of Canadian nursing homes, crowded homes were substantially more likely to be private for-profit facilities, as has been reported previously.^12^

Regulations, reimbursement schemes, and nursing home design standards have allowed crowding to persist, particularly in older homes. Medicaid provides no additional funds for private rooms, inadvertently disincentivizing construction of single-bed rooms. In the state of New York, design standards indicate that a minimum of only 10% of rooms need to be single bedded.^13^ In Canada’s largest province of Ontario, the design standards of 1999 and 2015 interdict construction of rooms with 3 or more residents, but sets no limits on the proportion of rooms with 2 beds, and 4 bed rooms were allowed to persist through a legacy clause.^14,15^ Recently a cap of 2-persons-per-room has been put into place for new admissions, but the occupancy cap is part of a directive specific for the COVID-19 pandemic, and as such, liable to be lifted.^6^ Consultations for new nursing home design standards in development in Canada indicate a strong preference for single-occupancy rooms.^16^

Evidence suggests that crowding is associated with increased SARS-CoV-2 incidence across a range of residential settings, including nursing homes, prisons and households.^4,17–20^ Our results extend these findings, suggesting that crowding is not only a risk factor for SARS-CoV-2 virus, but also associated with increased incidence of other acute respiratory infections. We hypothesize that crowded sleeping quarters have increased transmission across a range of mechanisms including aerosol, droplet, and direct and indirect contact. Regardless of the specific mechanisms of transmission, a reduction of crowding can be expected to reduce transmission rates since nursing home residents spend an average of 15 hours per day in bed (11 hours at night, and 30% of the remaining hours of the day).^21^ Furthermore, crowding impedes the ability to quarantine and self-isolate, which can only be partially mitigated by infection control measures.^22^

A systematic review on the burden of respiratory infection in nursing homes from the pre-COVID-19 pandemic period indicated that for nursing home settings, there was “little useful guidance for decision-making to decrease respiratory infection burden.”^23^ This lack of guidance was apparent during the COVID-19 pandemic, which had devastating impacts on nursing home residents in many countries. Our study identified an important and modifiable risk factor for non-COVID-19 respiratory infections and deaths in nursing homes. We found little heterogeneity across specific respiratory infections and consistency with a prior study of the association between crowding and SARS-CoV-2.^4^

There are additional benefits, as well as costs, associated with reducing crowding in nursing homes. Additional benefits include that most older adults prefer being housed in a single-bed rooms, by a wide margin.^24^ In addition, there are potential benefits in terms of reduced nighttime disturbances, though room sharing may not a primary driver of poor sleep among nursing home residents.^25^ The primary cost associated with decreasing crowding is financial, with estimated construction costs of single-bed rooms being 44% higher per bed compared to 2-bed rooms and 82% higher per bed compared to 4-bed rooms.^24^ Further, nursing staff in hospitals indicate that one perceived benefit of shared rooms are decreased walking distances during nursing shifts.^26^

Among limitations, our analysis was based on standardized outbreak surveillance procedures that may have missed some outbreaks and cases leading to errors in the total outbreak size, and may have misattributed the causal pathogen, as only the first 4 outbreak cases were tested in accordance with provincial guidance.^7^ Our analyses based on deaths may be less subject to misclassification errors and showed stronger associations with less heterogeneity. Data on occupancy, and resident and nursing home characteristics was extracted in January 2020, while nursing home respiratory outbreaks included in this analysis occurred between 2014 and 2019, meaning that we excluded all homes that closed before January 2020, and that the estimated home characteristics of residents may have changed. However, composition of nursing homes is extremely stable through time in Ontario; the median occupancy rate we observed, of 98.2% is consistent with that found in a 2012 auditor general report and the government strongly incentivizes homes to keep occupancy greater than 97%.^27^ The finding that 94% of outbreaks could be linked to a home further suggests few nursing homes closed during this time. Finally, an additional limitation of this work is that nursing homes with higher crowding index likely are also older, have smaller rooms, are more crowded in other common areas of the facility, and have lower ventilation rates, so that it may be difficult to attribute all differences to multi-occupancy rooms.

In this population-based study pre-dating the COVID-19 pandemic, crowded nursing homes experienced higher incidence of outbreak-associated respiratory infections and deaths, as well as outbreak frequency and size. This analysis can inform the decisions on the design and construction of nursing homes, and on the use of multi-bedded rooms in the future. Decreasing crowding in nursing homes is an important patient safety initiative beyond the COVID-19 pandemic.

## Data Availability

The data from this study is not publicly available.

